# PWAS Hub: Exploring Gene-Based Associations of Common Complex Diseases

**DOI:** 10.1101/2024.01.20.23300645

**Authors:** Guy Kelman, Roei Zucker, Nadav Brandes, Michal Linial

## Abstract

PWAS (Proteome-Wide Association Study) is an innovative genetic association approach that complements widely-used methods like GWAS (Genome-Wide Association Study). The PWAS platform involves consecutive phases. Initially, machine learning modeling and probabilistic considerations quantified the impact of genetic variants on protein-coding genes’ biochemical functions. Secondly, aggregating the variants per gene for each individual determines a gene-damaging score. Finally, standard statistical tests are activated in the case-control setting to yield statistically significant genes per phenotype. The PWAS Hub offers a user-friendly interface for an in-depth exploration of gene-disease associations from the UK Biobank (UKB). Results from PWAS cover 99 common diseases and conditions, each with over 10,000 diagnosed individuals per phenotype. Users can explore genes associated with these diseases, with separate analyses conducted for males and females. The PWAS Hub lists statistically significant genes associated with common diseases. It also indicates whether the analyzed damaged gene is associated with an increased or decreased risk. For each phenotype, the analyses account for sex-based genetic effects, inheritance modes (dominant and recessive), and the pleiotropic nature of associated genes. The PWAS Hub showcases its usefulness by navigating through such proteomic-genetic application for asthma. Graphical tools facilitate comparing genetic effects between the results of PWAS and coding GWAS, aiding in understanding the sex-specific genetic impact on common diseases. This adaptable platform is attractive for clinicians, researchers, and individuals interested in delving into gene-disease associations and sex-specific genetic effects. The PWAS Hub is accessible at http://pwas.huji.ac.il.

## Background

Public databases, such as the GWAS catalog, have compiled thousands of reports from genome-wide association studies (GWAS) (MacArthur et al. 2017). However, despite the abundance of GWAS, most studies have not significantly contributed to our understanding of disease mechanisms (Visscher et al. 2012; Brandes et al. 2022). The traditional GWAS approach faces challenges in mapping variants to their respective credible genes, leading to ambiguity and often unjustified variant-to-gene associations (Schaid et al. 2018). Notably, a large proportion of GWAS-associated variants (e.g., GWAS catalog, GWAS central) are located in regions with no functional annotations, such as intergenic regions and introns, making it difficult to gain meaningful insights into disease etiology (Mountjoy et al. 2021).

To address these gaps in contemporary genetic studies, we developed PWAS (Proteome-Wide Association Study), a method that conducts genetic association studies at the gene level (Brandes et al. 2020). PWAS aims to infer causal associations from variations within protein-coding genes. It utilizes a scoring method based on machine learning and probabilistic modeling to assess the functional effects of genetic variants and their impact on protein-coding genes (Brandes et al. 2019). The development of PWAS involved utilizing genotyping data from the UK Biobank (UKB) (Zhao et al. 2020). The input for PWAS includes all tagged and imputed SNPs within the coding regions of protein-coding genes. The underlying idea is that by aggregating weak effects across different variants, PWAS can detect genetic signals at the gene level. With a gene damage probability score per individual, robust and valid associations are identified by applying standard statistical methods. Specifically, linear regression models test associations for continuous phenotypes (e.g., BMI, height), while logistic regression models are used for binary phenotypes (e.g., schizophrenia, asthma). Notably, PWAS accommodates nonlinear genetic effects by considering dominant or recessive inheritance models for each associated gene (Brandes et al. 2019). PWAS complements GWAS and other gene-centric approaches like SKAT (Wu et al. 2011), providing more interpretable results by focusing on associations reliant on protein function alterations.

Traditionally, the sex differences in complex human traits were limited to physical and behavioral differences. It was attributed mainly to the influence of sex chromosomes and gene interaction with sex hormones. These differences are translated to anthropometric measurements (e.g., height, weight, adipose distribution) but to a lesser extent to human physiology and the mechanism of complex traits (Gilks et al. 2014). However, the exploration of sex-dependent genetics in human traits has become feasible in recent years due to the increased scale of resources with rich genomic and clinical data (Bernabeu et al. 2021).

The PWAS Hub leverages the results of PWAS for major diseases and conditions (e.g., asthma, hypertension, type 2 diabetes). It encompasses the human coding genome, encompassing over 18,000 protein-coding genes, with each gene tested according to dominant, recessive and combined inheritance models. PWAS Hub provides cross-references to relevant resources for genomics, proteomics, and phenotype-related information. It presents a user-friendly website with browsing capabilities for genes and medical outcomes. We conduct separate analyses for females, males, and both sexes, allowing interactive navigation across sexes and by the specific model of inheritance.

## PWAS Hub Construction

### Data Source

The input data for our study was sourced from the UK Biobank (UKB), a large database that encompasses detailed medical, genotyping, and lifestyle information for approximately 500,000 individuals aged 40 to 69 across the United Kingdom. The recruitment of participants took place between 2006 and 2010. To ensure consistency in our analysis, we focused on individuals of European origin (identified through data field codes 1, 1001, 1002, 1003, and information on ethnic background via data field 21000). In order to account for genetic relatedness, we considered genetic ancestry (referred to as genetic ethnic group, data field 22006) and randomly selected one representative from each kinship group while removing other genetic relatives.

The disease classification in our study relies on the International Classification of Diseases, Tenth Revision (ICD-10) codes for the diagnosis. Similar to other association studies, PWAS uses case-control settings. The tested cohort included individuals that had been diagnosed by the main or secondary codes (UKB data fields 41202 and 41204, respectively), and the summary diagnosis code 41270. The latter covers the distinct diagnosis codes a participant has recorded across all their hospital inpatient visits, in either the primary or secondary position. The filtered cohort has approximately 275,000 individuals of European origin. PWAS Hub covers diseases that are common and exceed 10,000 individuals in our filtered set who have been diagnosed with a specific disease or condition (i.e., cases). Each cohort carrying a disease phenotype was divided into three sets: males, females, and both. In specific cases, only one of the sexes was analyzed. For example, the ICD-10 C50 “malignant neoplasm of the breast” reported on 10,682 affected females, and only 77 affected males that were not analyzed.

### Variant inference by FIRM

The PWAS methodology assumes that variants in coding regions affect phenotypes by altering the biochemical function of the encoded protein of a gene. The FIRM (functional impact rating at the molecular level) is a pre-trained machine-learning (ML) model that estimates the extent of the damage caused to each protein for the entire proteome (Kelman et al. 2021). FIRM uses over 1100 numerical features to predict the effect score of any variant. For further details see https://github.com/nadavbra/firm. The performance of FIRM reached an AUC of 90% (precision = 86%, recall = 85.5%) and accuracy of 82.7% with respect to variants tagged as pathogenic/ likely pathogenic in ClinVar (Landrum et al. 2020). Each non-synonymous variant (e.g., missense, nonsense, frameshift, in-frame indel, and canonical splice-site variants) is assigned a functional effect score ranging from 0 to 1. Such a score captures the propensity of a variant to damage the gene’s protein product. The predicted effect score of a variant is normalized so that zero reflects a complete loss of function (i.e., LoF mutation) and 1.0 represents a protein with no effect on protein function (i.e., synonymous). Note that the calculated effect score is agnostic to the phenotype, as the score reflects the impact on function of the aggregated protein’s biochemical damage.

PWAS reports its results on ∼18,050 coding genes. The current list of a complete human proteome is extracted from UniProtKB (Boutet et al. 2016) with over 20,000 proteins. About 2000 of them are not included in the analysis of PWAS. Among these unmapped genes are genes that are characterized by ambiguous mapping of UniProtKB to RefSeq. Examples are endogenous retrovirus groups (e.g., ERVK). The gene list that is not covered by PWAS is referred to in the FAQ section http://pwas.huji.ac.il/FAQ#noGeneSymbol. It is also available for direct download under http://pwas.huji.ac.il/api/file/uniprot.not_mapped.tsv.gz

### Effect size of disease associations

Per-variant damage predictions are aggregated at the gene level for all listed variants. A protein functional effect score is reported for dominant or recessive inheritance modes. The dominant inheritance model only requires a single allele to be affected, while for recessive inheritance, both alleles should be mutated for gene functionality to be affected. These scores are between 1 and 0, where a value of 1 represents no predicted loss of functionality and no effect on the phenotype for either dominant or recessive modes. PWAS also provides a combined model for the protein-phenotype association that is fondly termed the “hybrid” model. This model uses both the dominant and recessive effect scores of each participant as covariates in a logistic regression over the phenotype. To find gene associations, PWAS tests for a correlation between the functional alterations and the phenotype of interest using standard case-control statistical methods. To determine the size of the effect of a gene on PWAS, we applied a statistical measure of Cohen’s d value. Cohen’s d value (standardized mean difference, SMD) is the normalized difference in the mean gene effect score between cases and controls. It was calculated independently for either dominant or recessive effect scores. In PWAS, positive values indicate a positive correlation with the gene effect scores, where higher effect scores represent less functional damage, thereby indicating the “protective” virtue of genes. To account for potential biases (e.g., batch effect in data collection, residual population structure, age), we included a collection of 172 covariates with: sex (binary), year of birth (numeric), 40 principal components of the genetic data that capture the ancestry stratification were provided by the UKB (numeric), the UKB genotyping batch (one-hot-encoding, 105 categories), and the UKB assessment centers associated with each sample (binary, 25 categories) (Zucker et al. 2023).

### Coding-GWAS analysis

While the PWAS effect scores use a gene-aggregation approach, we also performed a routine GWAS variant independent approach under a similar case-control design, while restricting the variants to the coding-region (coined coding GWAS, cGWAS). A total of 639,323 imputed variants (out of the 97,013,422 imputed variants) were included for analysis across the 18,053 protein-coding genes. Note that the set of variants used by PWAS and coding-GWAS is identical (Zucker et al. 2023). On average, there are 35.4 nonsense and missense mutations per coding gene.

The cGWAS associations was derived from PLINK 2.0 default logistic regression. The calculated z-score specifies the effect size and directionality of the effect per variant. Note that for GWAS, a positive z-score indicates a risk variant due to the positive correlation between the disease and the number of alternative alleles. To minimize bias in these complementary analyses, we also included the same 172 covariates that were used in PWAS. In the PWAS Hub portal, we report for each gene association the subset of significant variants from cGWAS that contributed to the association.

### Population analysis for a gene-centric risk

We tested the genetic signal of a disease in the entire population per gene. We examined the distribution of PWAS effect scores by considering all possible cutoffs (indicated by % of the cohort). For each gene-disease association study, we report the threshold that gave the best partition relative to the gene in the population presenting the disease in focus. For example, a threshold marked “bottom 10%” applies to effect-score values where the chosen threshold is 10% of the cohort with the lowest scores (i.e., the cohort decile with the greatest functional damage to the gene). Specifically, phenotype prevalence is calculated based on the 95% confidence interval using Wilson’s score for binomial proportion (Kelman et al. 2021). This best-cut analysis is performed for females, males, and both sexes. Each gene-disease association was tested under recessive or dominant inheritance modes.

### PWAS Hub Accessibility

The PWAS Hub portal presents an in-depth statistical analysis of findings according to the PWAS approach. Details on the underlying methodology are found in (Brandes et al. 2020). For additional support and to enhance the utility of the portal, we direct the user to ‘help and examples’. To learn more about its applicability, we invite the reader to browse through the tutorial (pwas.huji.ac.il/#tutorial-head), and FAQ (pwas.huji.ac.il/FAQ) sections of the portal. The PWAS Hub is available at http://pwas.huji.ac.il.

### PWAS Hub Application

The PWAS Hub portal covers 86 disease phenotypes based on ICD-10 diagnosis codes (**Table 1)**. This list can be viewed on the portal’s “Summary Page” http://pwas.huji.ac.il/service. Details on the ICD-10 terminology are provided via a direct link at the top panel of the “Summary Page”. Each phenotype was tested independently following a procedure of grouping the population into females, males, or both. The summary view highlights the genetic effects of major phenotypes that resulted in PWAS-associated genes for females and males.

**Table 1.**
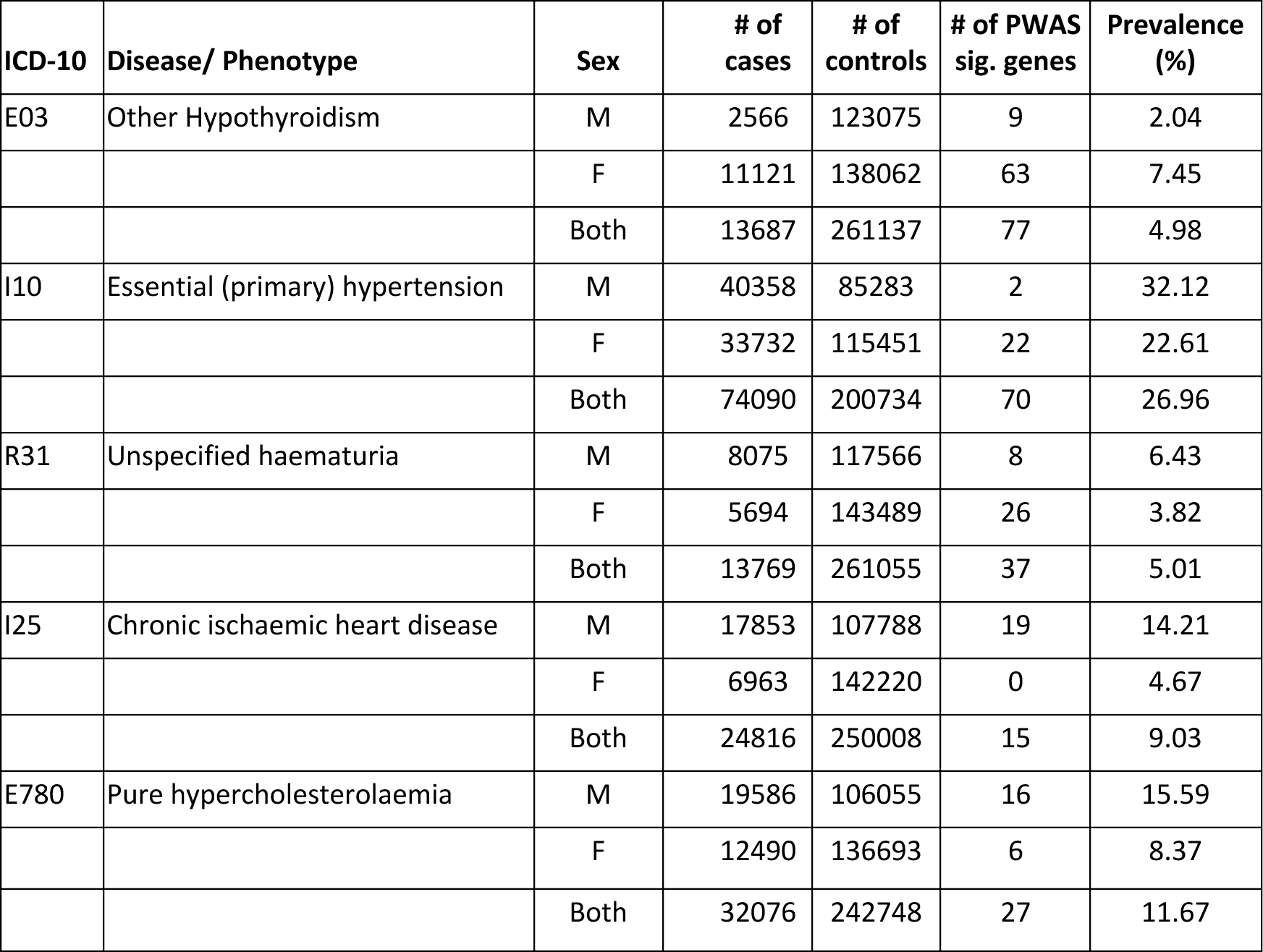
Samples of the Summary page from the PWAS Hub. Phenotypes are partitioned by sex.

| ICD-10 | Disease/ Phenotype | Sex | # of cases | # of controls | # of PWAS sig. genes | Prevalence (%) |
| --- | --- | --- | --- | --- | --- | --- |
| E03 | Other Hypothyroidism | M | 2566 | 123075 | 9 | 2.04 |
|  |  | F | 11121 | 138062 | 63 | 7.45 |
|  |  | Both | 13687 | 261137 | 77 | 4.98 |
| I10 | Essential (primary) hypertension | M | 40358 | 85283 | 2 | 32.12 |
|  |  | F | 33732 | 115451 | 22 | 22.61 |
|  |  | Both | 74090 | 200734 | 70 | 26.96 |
| R31 | Unspecified haematuria | M | 8075 | 117566 | 8 | 6.43 |
|  |  | F | 5694 | 143489 | 26 | 3.82 |
|  |  | Both | 13769 | 261055 | 37 | 5.01 |
| I25 | Chronic ischaemic heart disease | M | 17853 | 107788 | 19 | 14.21 |
|  |  | F | 6963 | 142220 | 0 | 4.67 |
|  |  | Both | 24816 | 250008 | 15 | 9.03 |
| E780 | Pure hypercholesterolaemia | M | 19586 | 106055 | 16 | 15.59 |
|  |  | F | 12490 | 136693 | 6 | 8.37 |
|  |  | Both | 32076 | 242748 | 27 | 11.67 |

**Table** 1 lists a sample of phenotypes partitioned into males, females, and both sexes. The number of associated genes is indicated. The total number of participants included in the PWAS Hub are divided to 149,183 females and 125,641 males The number of “both” (274,824) covers individuals of European origin where family related individuals were filtered out. Note that for some phenotypes, the prevalence of males and females is substantially different. Example is the hypothyroidism that shows a 3.6-fold higher occurrence in females versus males (**Table 1**). The full and current version of this table can be found at http://pwas.huji.ac.il/service.

### The PWAS Hub Layout

The welcome page of the PWAS Hub http://pwas.huji.ac.il summarizes the actions that can be performed. The user is thus presented with multiple actions in the form of a “What can I do here?” panel. The panel provides a brief overview of the portal utility. The user can select a gene of interest (GOI) as a starting point while seeking associations with any of the diseases listed in the Summary table (as in **Table 1)**. Alternatively, the user may choose a disease of interest (DOI) to estimate its gene-based associations and gain insights toward causality, disease mechanisms, and sex-dependent or independent genetic effects.

Figure 1 provides an operational workflow for the PWAS Hub. The portal highlights the centrality of sex-dependent genetics by beginning with the desired cohort of females, males, or both. At any stage, the user can compare the sex-based results by switching between the selected cohorts with the green highlighted (male/female) button. This button appears on any page that offers sex-based analyses. We introduce three main questions that the user may choose to address using the PWAS Hub: (a) focusing on a gene, what conditions is it associated with? (b) focusing on a phenotype, what are the genes that promote or lower its risk? and (c) given a phenotype, what would be the genetic differences across sexes, and consequently their sex-oriented risk factors?

**Figure 1.**
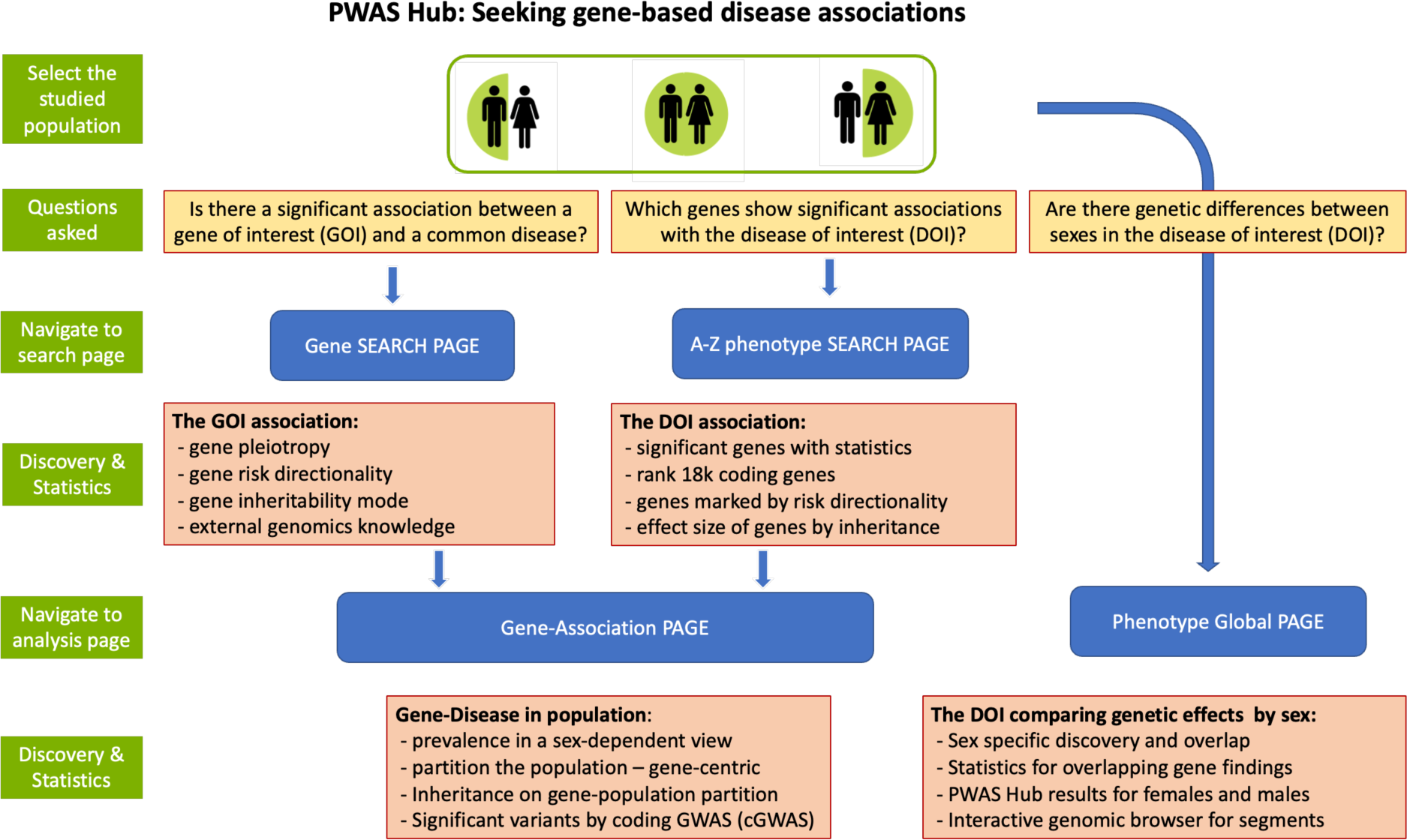
PWAS Hub layout. The primary option for activating the PWAS HUB portal is by selecting cohorts by sex. Then, three questions can be asked as described. The navigation to the “Search Page” and “Analysis Page” is shown. The main discoveries and supporting statistics are listed in the detailed orange frames associated with each of the major pages of PWAS Hub.

### PWAS Hub Implementation

For a full discussion on database schema and implementation, refer to the database page on PWAS Hub http://pwas.huji.ac.il/Database. **Supplemental Figure S1** shows the schema with constrained tables for the major portal displays. To appreciate the quantitative scale, the number of rows for each main table is shown. Specifically, the main PWAS disease-statistical table includes 5.1M rows, the variant-disease table comprises of 54.2M rows and the gene list include 18.1k, and gene variants table covers 225k informative rows.

### Navigating the asthma showcase in the PWAS hub

To illustrate the usability of the PWAS Hub as a navigation and discovery tool, we start by browsing the database using the “Phenotypes A to Z” search page. Figure 2 shows the layout of the resulting search for the letter A in the alphabetical index. The results of the “Phenotype A to Z” search allow an overview view of all PWAS gene associations, where for each phenotype, the number of significant genes is reported by colored symbols (acronym F for FDR statistics). Symbols report results by considering inheritance models (acronyms D, R, and H indicate dominant, recessive, and hybrid inherent modes, respectively). Summary statistics of the number of results according to the F, D, R, and H definitions are presented for each phenotype. Here, we showcase the flow for asthma (ICD-10: J45).

**Figure 2.**
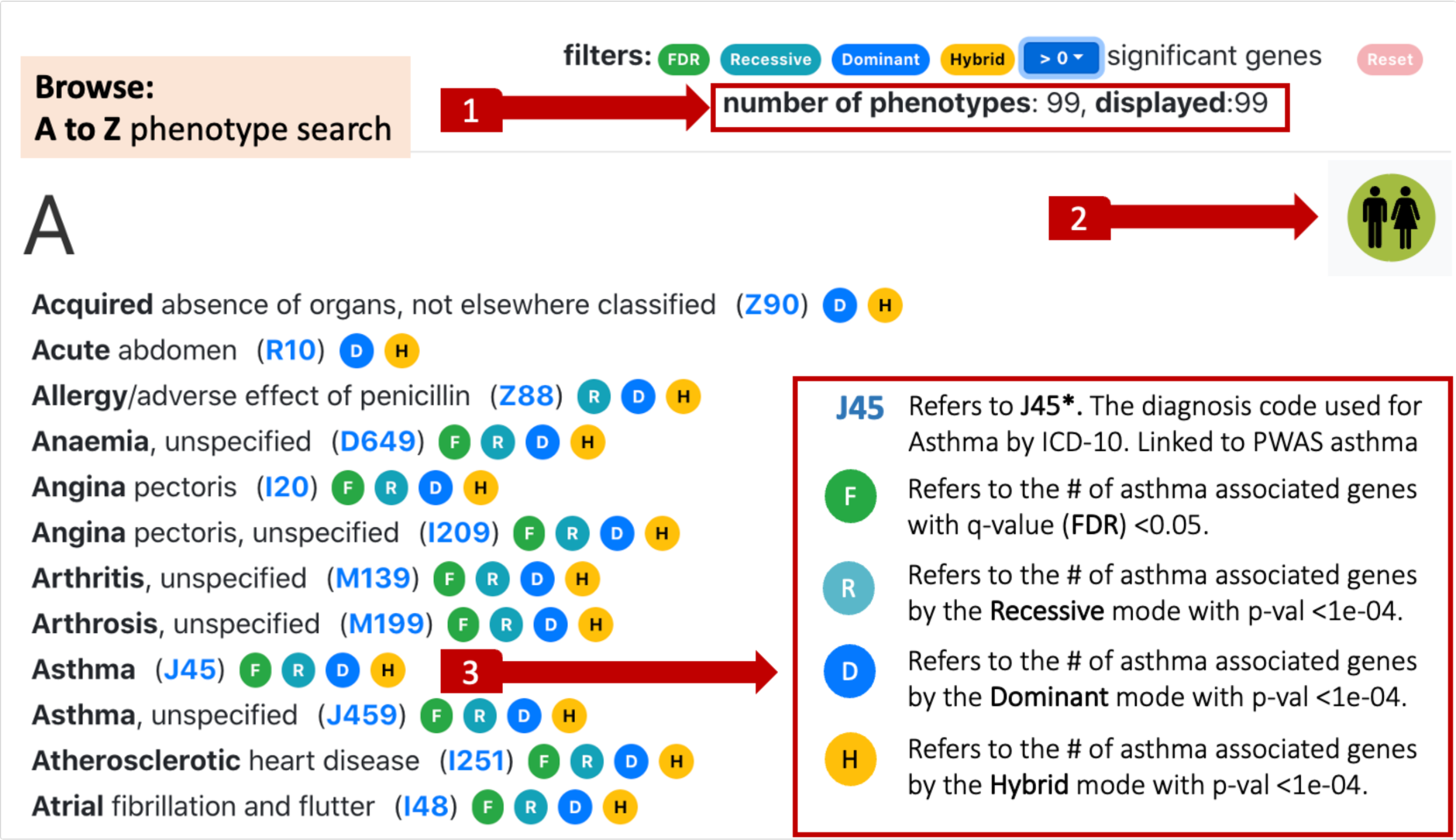
Browsing the “Phenotypes A to Z” search page at http://pwas.huji.ac.il/AZ. The user can filter out phenotypes by selecting a threshold for the number of significant results. Once filtered, **(1)** reports the subset size. The user selects whether the browsing and downstream analyses are restricted to females, males, or both. In this example, a cohort with both sexes was selected **(2)**. Each phenotype is indicated by its ICD-10 code (clickable links to the Phenotype Pages). Results that meet the preselected criteria are marked by the symbols abbreviated F, R, D, and H **(3)**.

The cohort selection (females, males, or both) is “sticky” in the sense that it is carried over to all pages. At any time, the user may switch to a different cohort by pressing the respective sex button on the top right.

Figure 3 gives the resulting top-10 genes identified for asthma. The gene-significant list includes 27 genes. We provide candidates with a relaxed threshold (q-value <0.1). PWAS reports on 42 asthma-associated genes meeting this threshold. Genes are colored to easily assess the genes that foster the greatest chance of increasing or lowering the risk. The range of colors marks Cohen’s d value. The red color indicates an increased risk, and the blue color represents positive Cohen’s d values for decreased risk. The effect sizes of the genes are coded by the color intensity according to the statistical significance (q-value). Associated genes can be sorted by Cohen’s d value with protective (i.e., reduced risk, colored blue with 11 of 27 genes) and the 16 genes indicating increased risk for asthma (red).

**Figure 3.**
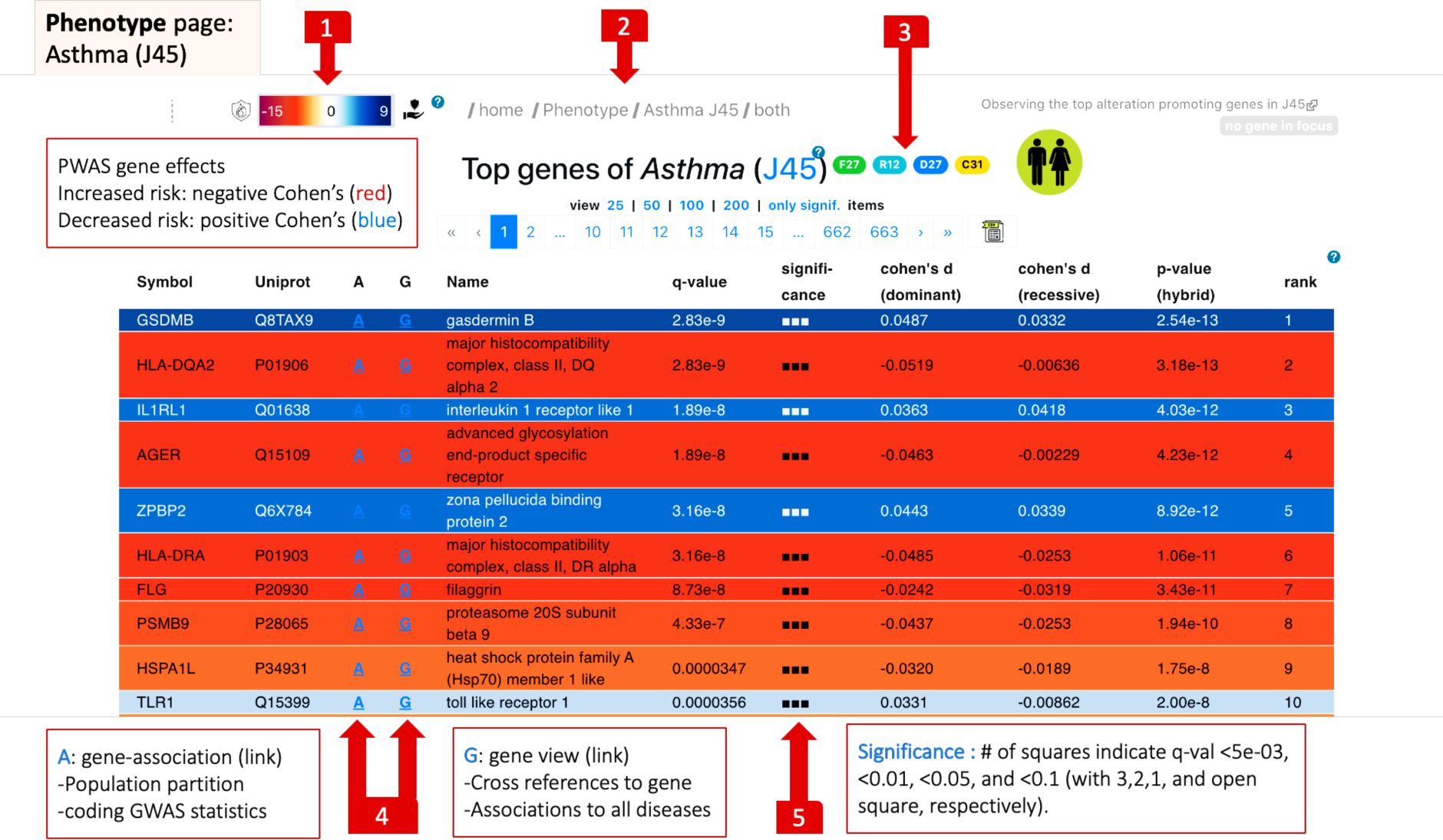
The phenotype page for J45 (asthma). The effect sizes of the genes are color-coded from red (negative Cohen’s d values = increased risk) to blue (positive Cohen’s d values = decreased risk) **(1)**. The intensity of the colors marks the statistical significance (q-value). Breadcrumb navigation links are available on all pages **(2)**. Summary statistics of the number of results according to the F, D, R, and H definitions, with an active link to the selected cohort. The link that marks J45 points the user to the “Phenotype Global” page **(3)**. Navigation to either the gene page (G) or the gene-association page (A) **(4)**. The significance of the listed genes is marked by a predetermined q-value symbolic scale **(5).**

Using the paging tool, the user may also choose to visualize either a significant set only (27 associated genes for asthma) or pages through lists of predetermined gene lists of 25, 50, 100, or 200 associated genes. Importantly, statistical analysis for each phenotype is available for all 18.1K genes. Each row in the table is linked to a navigation to the gene page (marked as G) or the gene-association page (marked as A, Figure 3).

To allow a flexible inspection of the gene association with asthma, the gene association table (Figure 3) is sortable by clicking on the headers. When resorting the table, the user can focus on significant genes that carry the inheritance signal of choice. The other sortable information concerns the effect size (Cohen’s d values), where the negative and positive values are associated with an increased or decreased risk, respectively. The top gene that was identified by PWAS for asthma is Gasdermin B (GSDMB). This gene is highly expressed in lung bronchial epithelium in asthmatic patients. Alteration the protein level of Gasdermin B indicates a reduced risk (colored blue, Figure 3). Interestingly, in animal model, an increased level GSDMB governs airway remodeling by increasing fibrosis in the absence of inflammation (Das et al. 2016).

To further investigate the relevance of the PWAS Hub to gain insights on asthma etiology, we navigate to the gene view of HLA-DQA2. It was ranked second in the significant gene list and the most significant gene for an increased risk (Figure 3, HLA-DQA2, red color). Figure 4 shows the information presented on the gene page. Note that one can navigate directly to gene-centric information by using the search options for the gene name (free text), gene symbol (as in GeneCards), UniProt or RefSeq ID, or by its genomic location.

**Figure 4.**
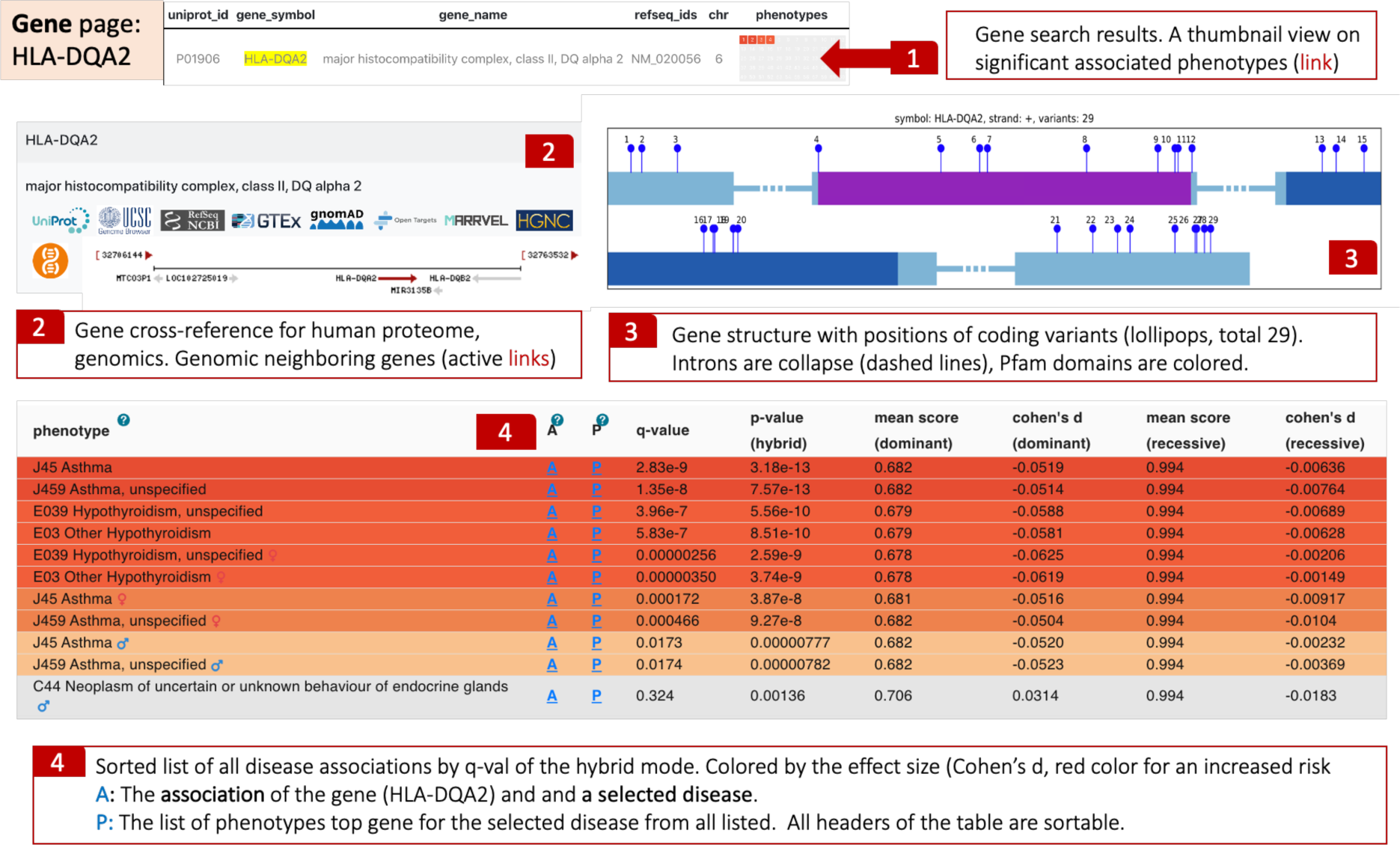
Gene-centric page. The search for a gene provides its protein and Refseq IDs and a thumbnail-colored view of the associated phenotypes. The red color depicts the effect size and directionality of an increased risk **(1).** Cross-references to major resources include an active map of the genomic location of the gene **(2)**. The numbered lollipops represent the coding variants that are included in the PWAS analysis, numbered along the protein length **(3)**. The table summarizes the statistics of this gene with all other phenotypes and diseases. The color depicts the effect size and directionality **(4).**

The gene page (Figure 4) provides a summary view of the statistical evidence for HLA-DQA2 and its association with any other listed diseases. We report on a strong association that is restricted to asthma and hypothyroidism, two immune-related common diseases. Importantly, gene associations are indicative of increased risk in both diseases, consistent with having a shared genetic basis. To further test the biological relevance of the subjected gene to asthma, the user can benefit from cross-reference resources. For example, the user can seek information on the gene association in the Open Target (OT) genetic portal. List of genomic variations from exomes and genome sequencing within a healthy population (e.g., gnomAD (Koch 2020)). Other public resources provide rich information on the gene, its protein product, and rich biological knowledge at molecular, cellular, and organismal levels (e.g., GeneCards, UniProKB).

While we demonstrate that the user can start with a gene of interest (GOI) or a disease of interest (DOI), the PWAS Hub application converges on the gene-phenotype analysis. For asthma, when seeking the associated gene list by sex, only nine genes were listed as significant in males. We illustrate the results for the phenotype gene for the PSMB9 gene (proteasome 20S subunit beta 9) that was ranked fourth in the male-dependent gene list. Figure 5 shows that the gene is highly significant for asthma in the hybrid mode (q-value FDR is 4.3e-07).

**Figure 5.**
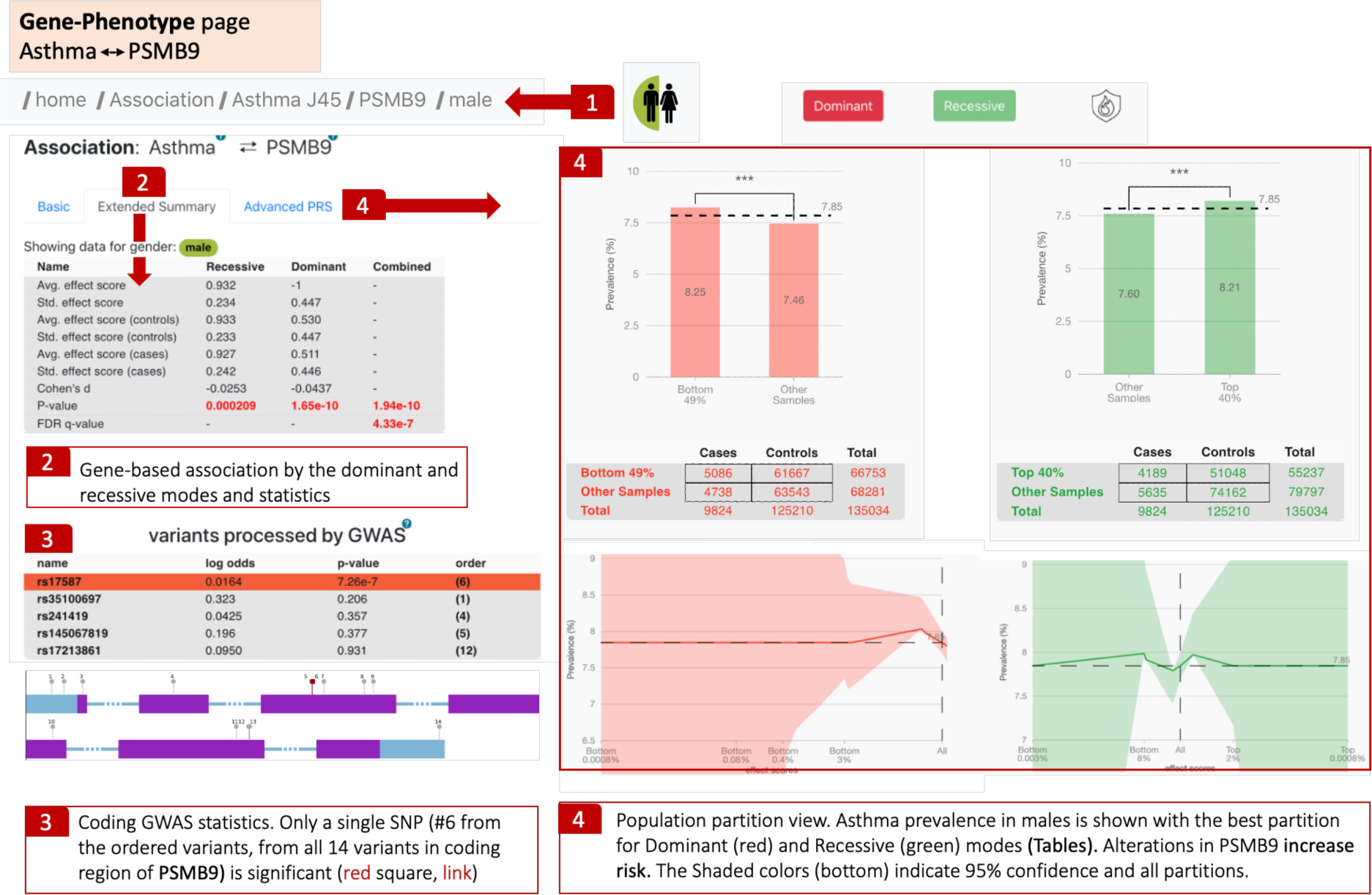
Gene-phenotype analysis page. The breadcrumb tool indicates the analysis is male-specific for asthma **(1)**. Extended statistical information is shared in a table with a calculated q-value for the hybrid heritability mode **(2)**. The results of coding-GWAS and the list of significant variants along the sequence. Each variant is labeled by its sequential number along with the coding exons. Significant variant (#6) for asthma is marked with a blue square **(3)**. The population partition view of dominant and recessive gene-population analysis is shown with 2×2 contingency tables used to determine the Fisher exact statistics (**4**).

An analytical view of the PSMB9 gene in the context of asthma is shown by the population partition view by gene (Figure 5). This view considers the partitioning of asthma-diagnosed individuals by the PWAS effect size of the gene. Specifically, we show (Figure 5) that by considering the dominant mode of PSMB9, in 49% of the population group (i.e., males) that have the lower score for the effect size, the prevalence of asthma is slightly higher. This is converted to an asthma prevalence of 8.25% (matched with 5086 male individuals with asthma), while the rest of the affected males have a lower prevalence of 7.46%. A similar trend was confirmed by considering the gene in recessive mode. This type of analysis refers to three main properties of the gene-disease associations: (i) what is the impact of modeling the gene by the different heritability modes; (ii) what percentage of the affected population can benefit from a simple population partition; and (iii) what is the gap in prevalence when the population is partitioned below and above such a cutoff partition line (i.e., for asthmatic males, it is 1.106).

Yet, in addition to the gene-based view, GWAS Hub displays the underlying variants within the coding gene. In the case of PSMB9, while the coding region of the gene has 14 variants (Figure 5), only a single SNP (rs17587) is statistically significant for the association with asthma. The SNP is very rare in gnomAD (apparently a healthy population), with a minor allele frequency (MAF) of 1.4e-05. This SNP was proposed to increase the risk of rheumatoid arthritis (RA) for the ethnicity of Han Chinese from Yunnan (Yu et al. 2013) and vitiligo in the Saudi community (Mufti et al. 2021). These observations argue for shared mechanisms in asthma and other autoimmune diseases.

Finally, we present a global view that allows a direct comparison of the genetic effects on gene-phenotype associations by sex. To illustrate this, we compared the genes identified by females and males for asthma. Figure 6 shows that the analysis by PWAS resulted in 9 significant genes for males and 9 genes for females. The indication is that there are no female-specific identified genes, but two genes are only identified in males. These male-exclusive genes are IL6R and MAP9.

**Figure 6.**
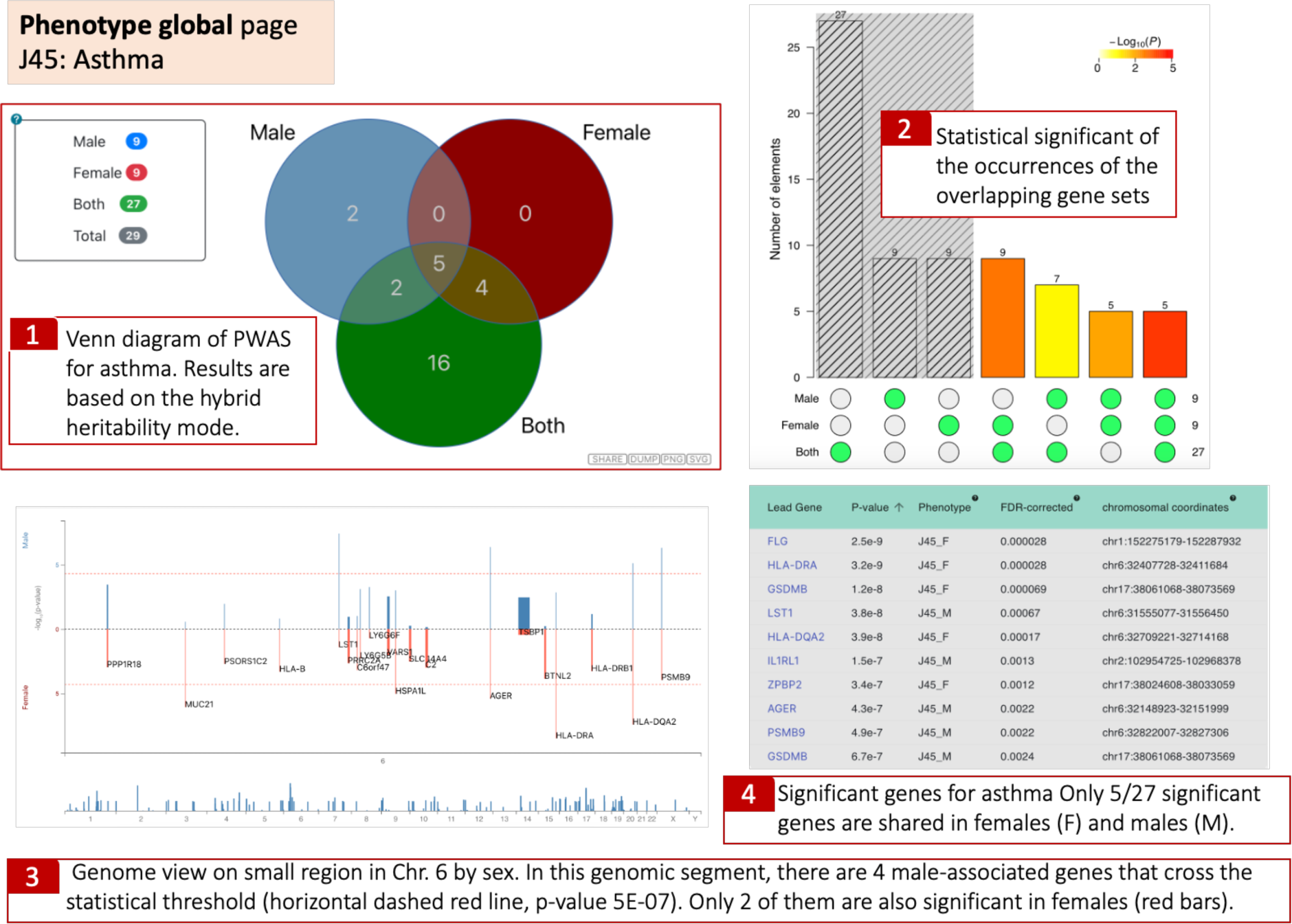
A collapsed view in the “Phenotype global page” with a comparison of the genetic effects by sex. The Venn diagram shows the results for males, females, and both with the number of genes identified by PWAS Hub (**1**). A visual of a three-way Fisher’s exact test is presented by the upset plot The grey shaded area is for the unions among the three subsets, thus there is no joint occurrence to estimate. The p-values are estimate to the p-value of having the exact subset of genes with respect to a random occurrence **(2).** Manhattan-like plot that maps the identified genes on the genome. The y-axis gives significance values for the enrichment for males (blue) and females (red). The x-axis depicts the chromosomal coordinates. A zoom in option is implemented from a genomic overview. More information is available by popup options with hover over a gene name **(3)**. A sortable summary table for each gene is shown for all gene according to their sex stratified asthma (J45) **(4)**.

Importantly, IL6 signaling through its receptor (IL6R) was associated with sex stratification as a risk factor for several autoimmune diseases (Hong et al. 2021). However, testing specifically asthmatic and healthy young males failed to meet statistics with regard to IL6R (Rantala et al. 2011). More surprising is the finding that MAP9 is higher in males. There are three informative variants (out of 57 along the coding region) in males, and none in females. No information was reported regarding MAP9 polymorphism or expression with respect to asthma.

Interestingly, most of the signals in females and males are located in the MHC locus on Chr. 6, which covers immunological risk factors for autoimmunity and many immune-related diseases. PWAS Hub provides an entry point for interactively searching sex-specific results across the genome (Figure 6). The genes are displayed by their genetic effects for males and females along the chromosomes using an interactive two-way Manhattan plot. We illustrate a snapshot of the genome browser for a small section of the MHC within Chr. 6. There are 4 genes that are significant in males in this genomic segment, only two in females, with only HLA-DQA2 (discussed in Figure 4) are significant in each sex. The graphical results are presented along with a detailed tabular list the sex-dependent informative genes.

## Discussion

PWAS (proteome-wide association study) detects gene-phenotype associations through the effect of variants on protein function, thus enhancing interpretability for complex diseases (Brandes et al. 2020). In the current version of the PWAS Hub, we discuss diseases according to clinical diagnosis codes based on ICD-10 indices. The results presented in PWAS Hub address the major pitfalls and limitations of GWAS, as discussed in the case of primary hypertension (Zucker et al. 2023) or predisposition to 10 cancer types in a large population (Brandes et al. 2021). In the implementation discussed in the PWAS Hub, we observed a relatively low number of significant genes for the studied diseases (e.g., 27 genes for asthma and 29 unique genes for females, males, and both) (Fig. 6). For example, for Angina pectoris (ICD-10: I20) only 3 genes were identified as significant for males and none for females. For only 16 diseases, the significant gene list includes >10 genes (Summary page, Table 1). The discovery of PWAS relies on a routine gene-centric statistical analysis. Specifically, in the last phase of PWAS, a statistical test is performed by comparing the mean and standard derivation of the partition to cases and controls per each of the heritable gene models. Therefore, irrespective of whether the PWAS score reflects the true underlying biology or not, it is guaranteed to minimize type I errors (false positive hits). Furthermore, as opposed to the variant additivity consideration used by GWAS, the aggregating scheme in PWAS considers heritability modes with the notion of recessive or dominant modes. We confirmed that in the case of cancer’s predisposition, the contribution of recessive inheritance is far larger than anticipated (Brandes et al. 2021).

By illustrating the results of PWAS Hub for asthma, we noted that many associated gene were immune-related, and specifically associated with autoimmunity (e.g., mostly occurring within the MHC locus). However, such trend was not replicated across phenotypes and was exclusively identified in disease with strong immune basis (e.g., psoriasis, type 1 diabetes, hypothyroidism) (Brandes et al. 2020). While discussing the results for disease mechanism is beyond the scope of this database paper, the list of discovered genes suggests a high level of specificity.

The current PWAS Hub focuses on 99 common complex diseases with an artificial definition of at least 10,000 cases from the UKB population (restricted to European origin, with no family relatedness). It is likely that medical records for people who have developed any of these common diseases are not updated. Therefore, some records are noisy and probably attenuates the success of true discovery. We will expand the utility of the PWAS Hub platform in the future. We plan to cover complex diseases according to PheWAS mapping (Denny et al. 2010). Under this classification, we have collected about 700 phenotypes (defined by PheCode) with a minimum threshold of 200 cases per PheCode. We will also expand the PWAS Hub to include quantitative and categorized traits (e.g., measurement from blood tests, diastolic pressure). Note that the foundation of PWAS was developed to deal with continuous and binary traits (Brandes et al. 2020). Finally, we expect to increase the discovery power by including rare and ultra-rare variants in the PWAS scoring methods. UKB provides a complete set of exome sequencing results that can be used in gene-based association methods (Karczewski et al. 2022).

### Conclusions

The PWAS score operates on an aggregated gene level, consolidating results that provide a quality gene list. This approach enhances interpretability, allowing for better assessment of causality and functional implications. We anticipate that the inclusion of personalized exome sequencing, even if the rare and ultra-rare SNVs occur infrequently, those with relatively high effect sizes will enhance discoveries without affecting gene-based statistical analyses. We propose that examining and navigating through various pages and inter connected perspectives offered by the PWAS Hub, can have substantial implications for personalized treatment strategies and improved disease management. We conclude that focusing on sex stratification is a valuable approach to highlight sex related disease etiology.

### Summary for Availability and Requirements

- Project Name: PWAS Hub
- Project homepage: http://pwas.huji.ac.il
- Operating system: Ubuntu Linux
- Programming language: Python, javascript
- Restrictions for non-academics: No license needed

### Abbreviations

DOI, Disease of interest

GOI, Gene of interest

GWAS, Genome-wide association studies

PheWAS, phenome-wide association study

PWAS, Proteome-wide association studies

MAF, Minor allele frequency

SNP, Single nucleotide polymorphism

SNV, Single nucleotide variant

UKB, UK biobank

WGS, Whole-genome sequencing

## Declarations

The authors declare that they have no conflict of interest. No competing financial interests in relation to the work described.

## Ethical approval

The study was approved by the University Committee for the Use of Human Subjects in Research Approval number 12072022 (July 2023). This study uses the UK-Biobank (UKB) application ID 26664 (Linial lab).

## Data Availability

All data produced in the present work are contained in the manuscript

http://pwas.huji.ac.il

---

**Supplementary Figure S1.**
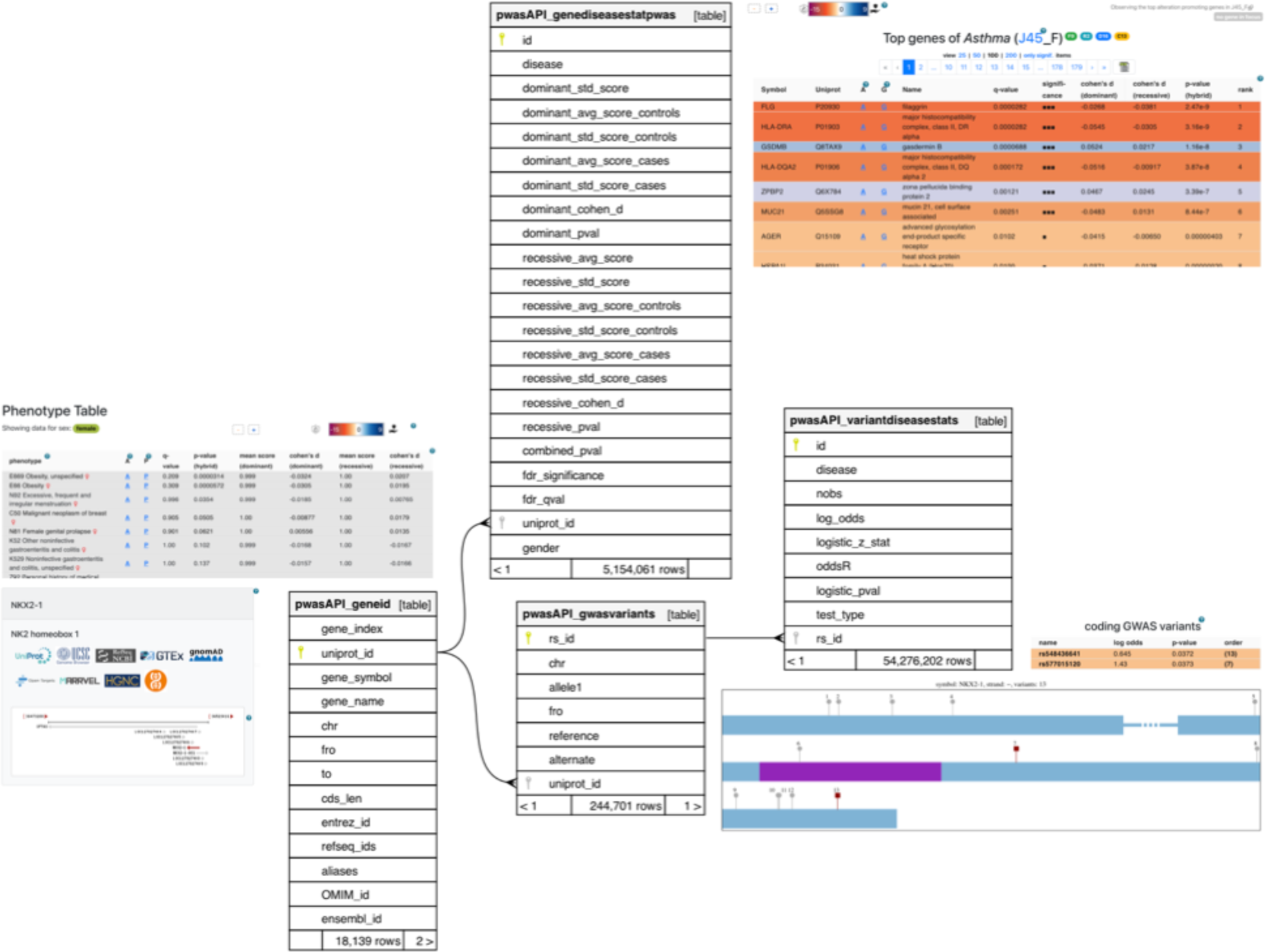
PWAS constrained tables.

**Table S1.** At the side of each table description appears the main visual that draws information from that table. *pwasAPI_geneid* connects all the tables that require gene information such as genomic coordinates, and the gene’s name in multiple vocabularies. *pwasAPI_genediseasestatpwas* is the main gene-disease correspondence that gives the statistics of the association pattern. The other tables store variant-level information.

